# Comparative Risk of Stroke Associated with GLP-1 Receptor Agonists and SGLT2 Inhibitors in Veterans with Type 2 Diabetes

**DOI:** 10.64898/2026.05.13.26353028

**Authors:** Sophia C. Sun, Serena C. Houghton, Yanping Li, Xuan-Mai Nguyen, Kelly Cho, Hugo J. Aparicio, Peter W.F. Wilson

## Abstract

**Introduction:** Stroke is a leading cause of disability and death in adults with type 2 diabetes (T2D). We evaluated the comparative stroke risk in Veterans with T2D initiated on either of two glucose-lowering medications: GLP-1 receptor agonists (GLP-1RA) or SGLT-2 inhibitors (SGLT2i).

**Patients and Methods:** We conducted a retrospective cohort study on diabetic Veterans aged 40 and older with no prior history of stroke or transient ischemic attack, who started on a GLP-1RA or SGLT2i between 2014 and 2021. Patients with contraindications or prior exposure to medication were excluded. Using national Veteran health data, we identified 195,072 eligible individuals and followed them from treatment initiation until stroke, death, loss to follow up, or end of follow up, whichever came first. Primary outcome was incident stroke, and secondary outcomes included ischemic and hemorrhagic stroke. We applied Kaplan-Meier methods and Cox proportional hazards models. Adjusted associations were estimated using inverse probability weighting.

**Results:** Both unadjusted and adjusted analyses suggest GLP-1RA users have reduced stroke incidence compared SGLT-2i users (HR = 0.67, 95%CI 0.64-0.69;HR = 0.72, 95%CI 0.69-0.75). Similar results were found in secondary outcome and stratified analyses, with GLP-1RA users having reduced stroke risk compared to SGLT2i users for all age groups, chronic kidney disease stages, and hemoglobin A1c levels.

**Discussion and Conclusion:** GLP-1RA treatment was associated with a lower risk of stroke compared with SGLT2i treatment in Veterans with T2D. These findings were consistent for ischemic and hemorrhagic strokes, suggesting potential differences in stroke risk between the treatments.

## Introduction

Type 2 diabetes is an independent risk factor for stroke and is prevalent among the US Veteran population, contributing to overall health and mortality rates. Individuals with diabetes are at a high risk of developing cardiovascular disease (CVD) and its associated and mortality ^1^.

Stroke also remains a major cause of morbidity and mortality. Within the Veterans Affairs healthcare system, the prevalence of acute ischemic stroke (AIS)—which accounts for most stroke cases—has increased steadily from 2006 to 2018, reaching approximately 120 cases per 1,000 Veterans, with an incidence of approximately 14 per 1,000 Veterans in 2018 ^2^.

Glucagon-like peptide-1 receptor agonists (GLP-1RA) are primarily used in treating type 2 diabetes (T2D) by lowering serum glucose levels to regulate metabolism ^3^. Similarly, sodium-glucose cotransporter-2 inhibitor (SGLT2i) are prescribed to improve glucose lowering capabilities in patients, in additional to providing cardiovascular and renal protein ^4^. SGLT2i medication usage became in the VA in 2013, while GLP-1RA usage began earlier in 2005.

With higher CVD burden in the Veteran population, this raises the question of the comparative effects of GLP-1RA and SGLT-2i usage in reducing stroke risk. These medications have shown to reduce major adverse cardiovascular events (MACE) and cardiovascular mortality in comparison to placebo ^5^. In recent years, use of GLP-1RA and SGLT2i medications is increasing, but there is still a lack of direct comparisons between the two drugs and their effects on incident stroke.

This study aims to address this gap by conducting a retrospective cohort analysis to compare the incidence of first stroke in patients using GLP-1RA versus SGLT2i treatment. Specifically, we assess the comparative effectiveness of these two drug classes on general stroke incidence and stroke subtype incidence, including acute ischemic stroke (AIS), intracerebral hemorrhage (ICH), and subarachnoid hemorrhage (SAH).

### Patients and Methods

#### Data Sources

This retrospective cohort study used clinical data from three national sources: (1) the Veterans Health Administration (VHA) Corporate Data Warehouse (CDW), accessed through the Veterans Informatics and Computing Infrastructure (VINCI);(2) Centers for Medicare &Medicaid Services (CMS);and (3) mortality data from the National Death Index (NDI). The study period began in 2014 to coincide with the availability and clinical usage of SGLT2i, given that GLP-1RA treatments were being used in the VA prior to this time. The CDW provided information on demographics, diagnoses, laboratory values, vitals, outpatient and inpatient encounters, and prescription records. CMS data supplemented utilization and diagnostic information for Veterans receiving both VA–Medicare care. NDI data were used to confirm death and classify stroke-related mortality (Department of Veterans Affairs, 2023). The study was approved by the Emory University Institutional Review Board and the Atlanta VA Medical Center Research &Development Committee under an informed consent waiver.

#### Study Population

The cohort included adults aged ≥40 years with type 2 diabetes who began usage of a GLP-1RA and/or SGLT2i during the study period between January 1, 2014, and December 31, 2021. Patients were required to have at least one type 2 diabetes diagnosis code (ICD9 250.x and ICD10 E08-13) and an active antidiabetic medication prescription prior to baseline date. Exclusion criteria included patients with a stroke or transient ischemic attack (TIA) prior to index date. TIA was defined with ICD-9 435.x and ICD-10 G45.x (excluding G45.4). There were no sex-based exclusions. The index date or baseline was defined as the date of first prescription to either a GLP-1RA or a SGLT2i after January 1^st^, 2014.

#### Exposure Definition

The exposure of interest was first use of either a GLP-1RA or an SGLT2i. GLP1-RA medications included semaglutide, dulaglutide, liraglutide, tirzepatide, exenatide, albiglutide, and lixisenatide. SGLT2i medications included empagliflozin, canaglifozin, dapagliflozin, and ertuglifozin. First use was defined for patients with no prior use of either drug class preceding the index date. For individuals who initiated both classes during follow-up, the class first prescribed at least a year prior was considered as the primary exposure. Medication initiation and dispensing records were identified using standardized VHA CDW data.

#### Outcome Ascertainment

The primary outcome was incident stroke, defined as a new stroke diagnosis occurring after index date, identified using ICD-9 and ICD-10 codes from both inpatient and outpatient settings using VA and CMS data. Secondary outcomes included ischemic and hemorrhagic stroke. Prevalent stroke was defined as stroke diagnosis occurring prior to index date, and these individuals were excluded. A stroke event defined as patients having had either ≥1 inpatient diagnosis code or ≥2 outpatient diagnosis codes for stroke.

AIS was identified using ICD-9 codes 433.x1, 434.x (excluding 434.x0), and 436.x, and ICD-10 code I63.x. Hemorrhagic stroke was defined as occurrence of either ICH or SAH. ICH was defined by ICD-9 431.x and ICD-10 I61.x, and SAH by ICD-9 430.x and ICD-10 I60.x. Deaths attributable to stroke were confirmed using NDI codes.

#### Covariates for analyses

Baseline covariates included: age at index date, sex, and race/ethnicity. The closest laboratory measures to the index date within 12 months prior were assessed: systolic and diastolic blood pressure, body mass index (BMI), hemoglobin A1c (HbA1c), total cholesterol, high density lipoprotein, and estimated glomerular filtration rate (eGFR). Smoking status and comorbidities such as coronary artery disease (CAD), heart failure (HF), cardiovascular disease (CVD), atrial fibrillation (AF), chronic kidney disease (CKD), prior myocardial infarction, and other vascular risk factors at any time prior to index date were recorded. CVD was defined as incidence of both CAD and HF. Comorbidities were defined using ICD-9 and ICD-10 codes, except for CKD. CKD was defined upon individuals having two eGFR values <60 mL/min/1.73m^2^, one taken closest to index date and one ≥90 days prior to index date. Medication covariates included statins and antihypertensives prescribed prior to index date.

### Statistical Analyses

Analyses were conducted using R (version 4.5.2). Patients were followed from index date until first stroke event, death, loss to follow-up, or baseline date. Inverse probability weights (IPW) were used to adjust for the probability of initiating either medication based on baseline covariates of age, gender, race/ethnicity, BMI, antihypertensive usage, statin usage, smoking status, acute pancreatitis (AP), CKD stage, HbA1c, AF, CKD, HF, CAD, CVD, and systolic blood pressure. AP was defined as ICD-9 code 577.0 and ICD-10 code K85.x. Kaplan–Meier curves compared time to first stroke across exposure groups. Cox proportional hazards models incorporating IPW were used to estimate hazard ratios (HRs) and 95%confidence intervals (CIs) comparing incident stroke occurrence between treatments.

### Sensitivity Analyses

Sensitivity analyses were conducted to evaluate the findings across several demographics and clinical factors. Analyses include stratification by age, gender, race/ethnicity, BMI, HbA1c level, and CKD stage. Age was stratified by those <65 years old and ≥65 years old (age eligible for Medicare). Race and ethnicity analyses were stratified into White, non-Hispanic;Black, non-Hispanic;Hispanic;American Indian/Alaska Native;Native Hawaiian/other Pacific Islander;Asian;and Unknown. CKD stages were categorized by guidelines using eGFR values: stage 1 (≥90 mL/min/1.73m^2^), stage 2 (60–89), stage 3 (30–59), stage 4 (15–29), and stage 5 (<15). Stage 4 and Stage 5 were combined for analytical purposes. BMI was characterized by national categorization guidelines: underweight (<18.5 kg/m^2^), normal (18.5-25), overweight (25-30), and obese (≥30). HbA1C level was stratified based off lab values: normal (<5.7%), pre-diabetic (5.7-6.4%), and diabetic (>6.4%).

An additional sensitivity analysis using a time-varying model was conducted to consider individuals who stopped or switched treatments during follow-up time. Cox proportional hazard models with IPW were used to estimate HRs and CIs, accounting for changes in treatment use.

## Results

193,347 patients were included in the study as users of a GLP-1RA or an SGLT2i medication (Figure 1). 206,349 Veterans with T2D initiated a GLP-1RA or an SGLT2i medication during the interval between our index and end date. 7,077 were excluded due to missing blood pressure, BMI, HbA1c, and eGFR data. 5,295 were excluded for having a stroke or TIA prior to their baseline date. Over a median follow-up time of 1.96 years, there were 12,595 (6.5%) strokes, of which 12,277 were ischemic and 318 were hemorrhagic.

**Figure 1.**
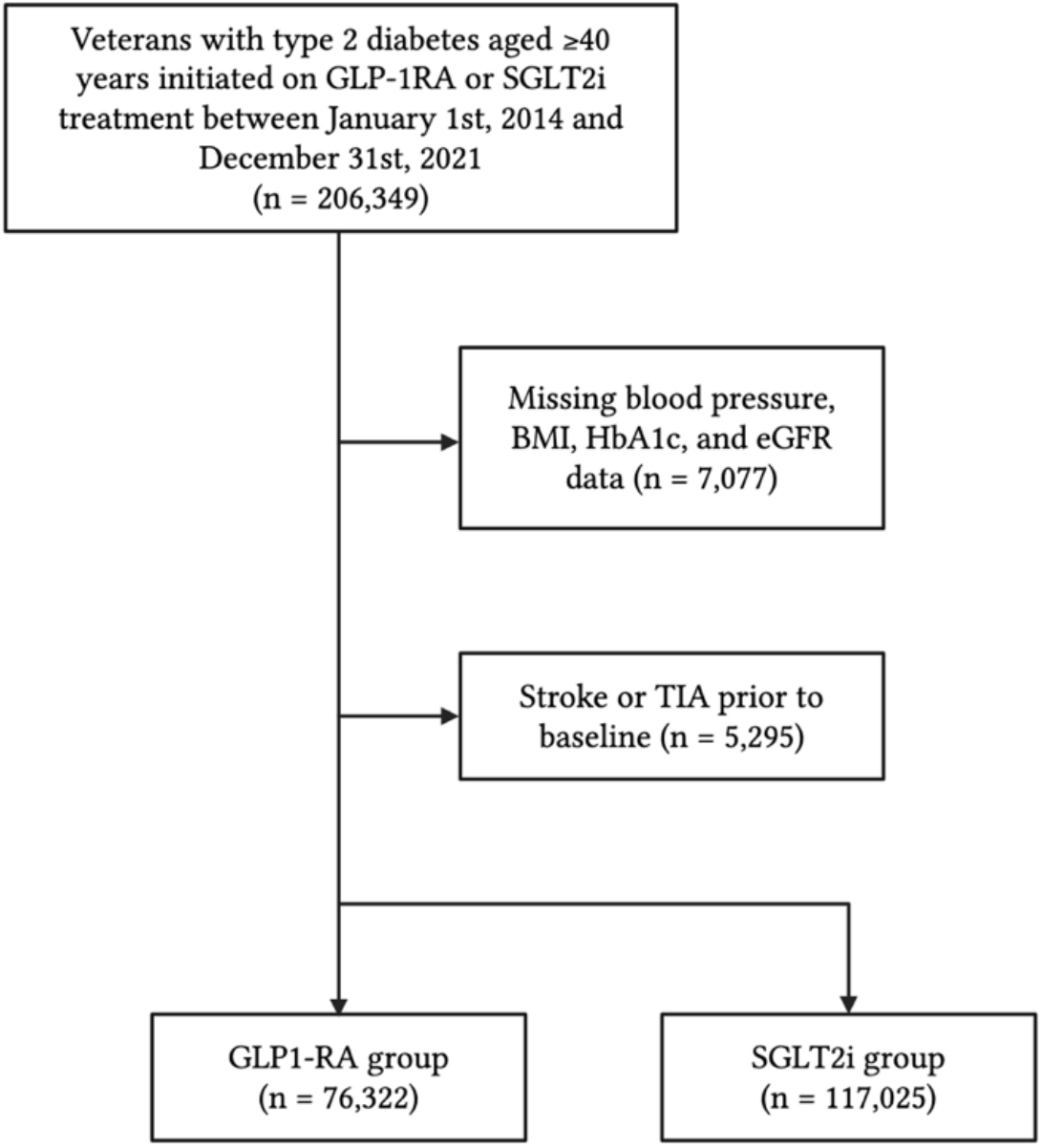
Flowchart of subject inclusion.

**Figure 2.**
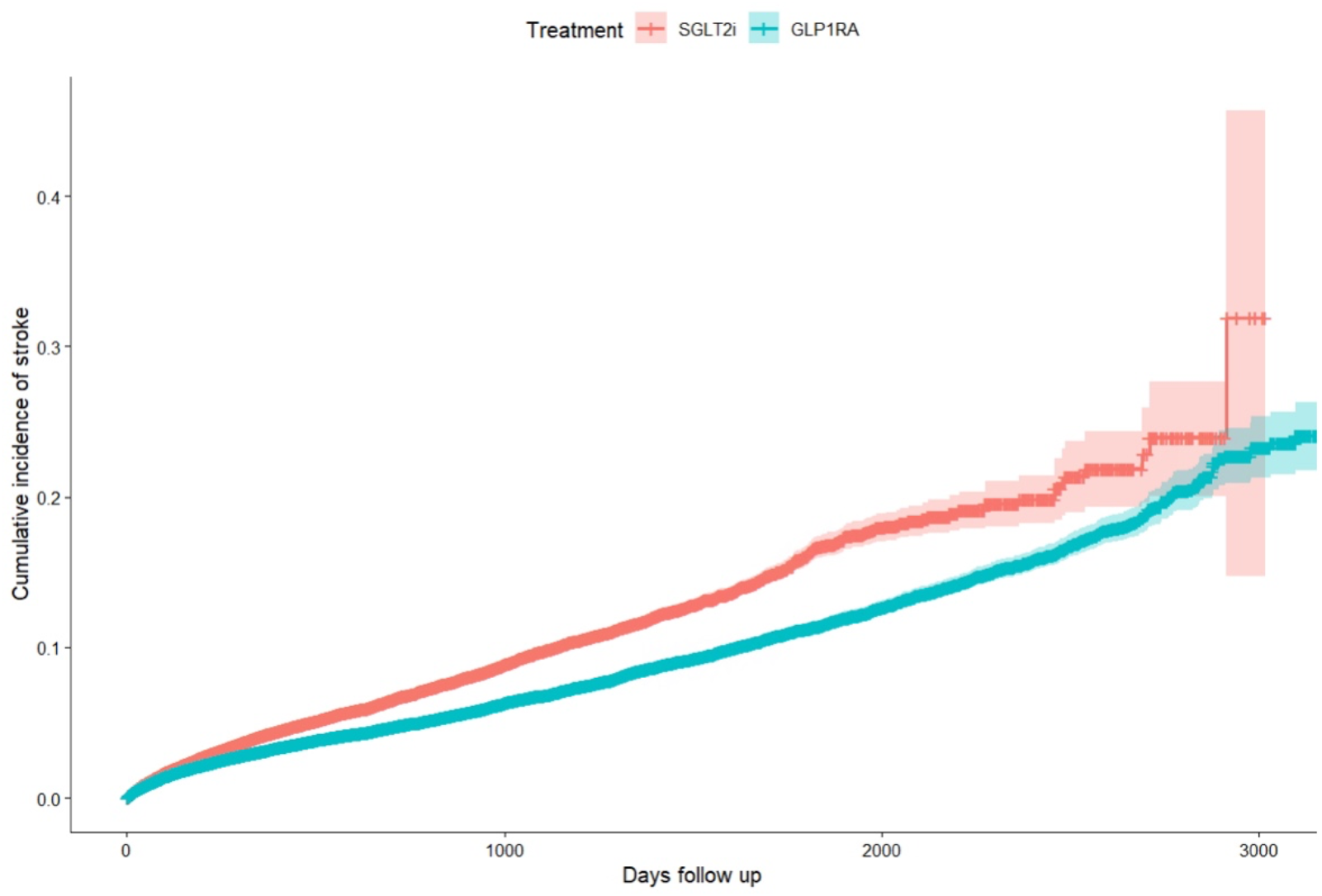
Adjusted cumulative incidence of stroke among new users of GLP-1RA vs SGLT2i treatments. Red line indicates cumulative incidence for SGLT2i users;blue line indicates cumulative incidence for GLP-1RA users. Shaded regions represent 95%confidence intervals. Data is adjusted using inverse probability weighting to account for patient age, gender, race/ethnicity, BMI, antihypertensive usage, statin usage, CKD status, smoking status, AP status, and CKD stage.

Baseline characteristics for the cohort are presented in Table 1. The mean age was 67.2, 95.5%were men, and the mean BMI was 33.9. Compared with SGLT2i users, GLP-1RA users were younger (median 66.4 vs 67.7), more likely to be female (6.2%vs 3.4%), and more likely to have a higher BMI (median, 35.6 vs 32.8). All participants had diabetes and were using diabetes medication at baseline. The median follow-up time for GLP-1RA and SGLT-2i users was 2.75 and 1.51 years, respectively.

**Table 1.**
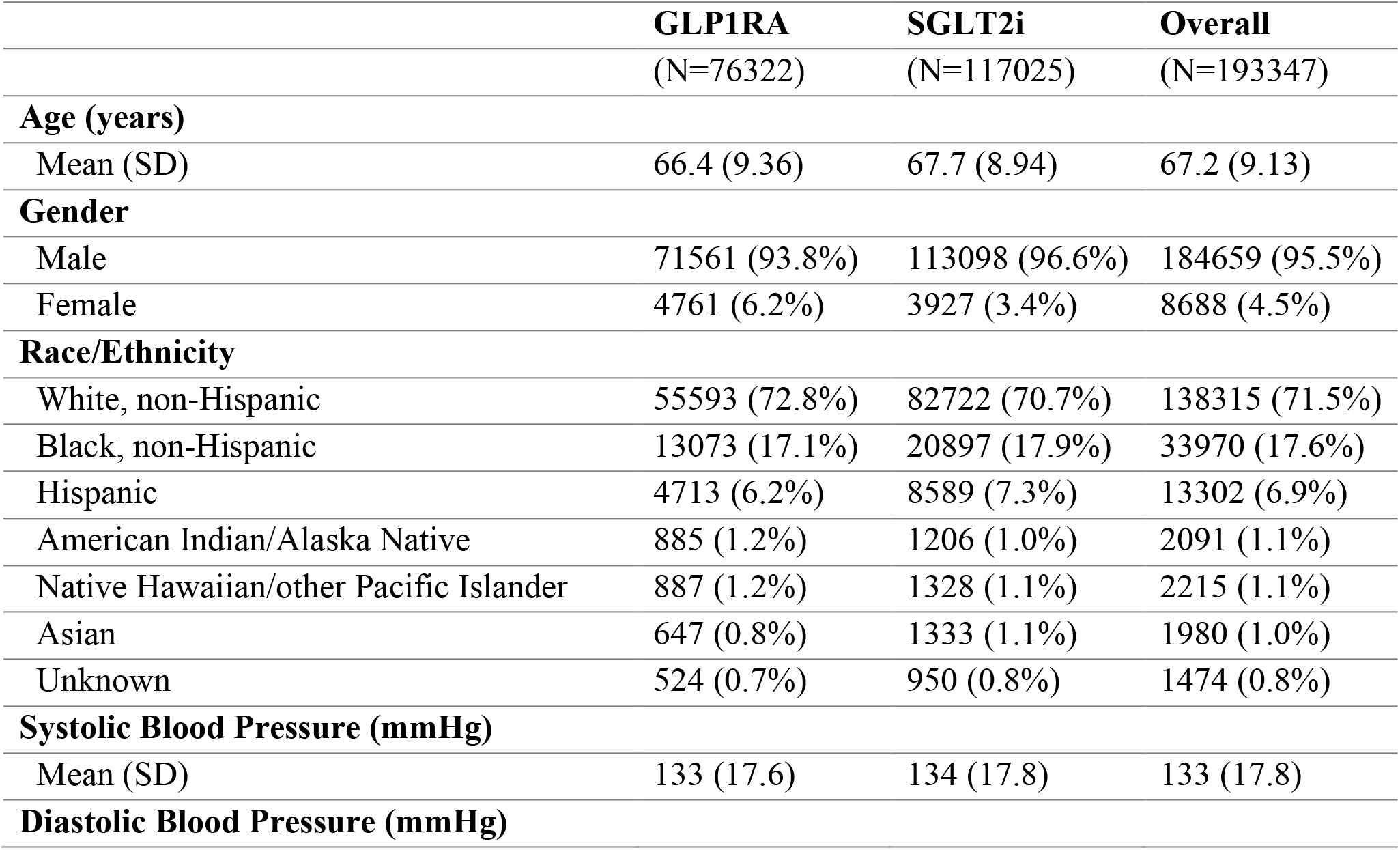
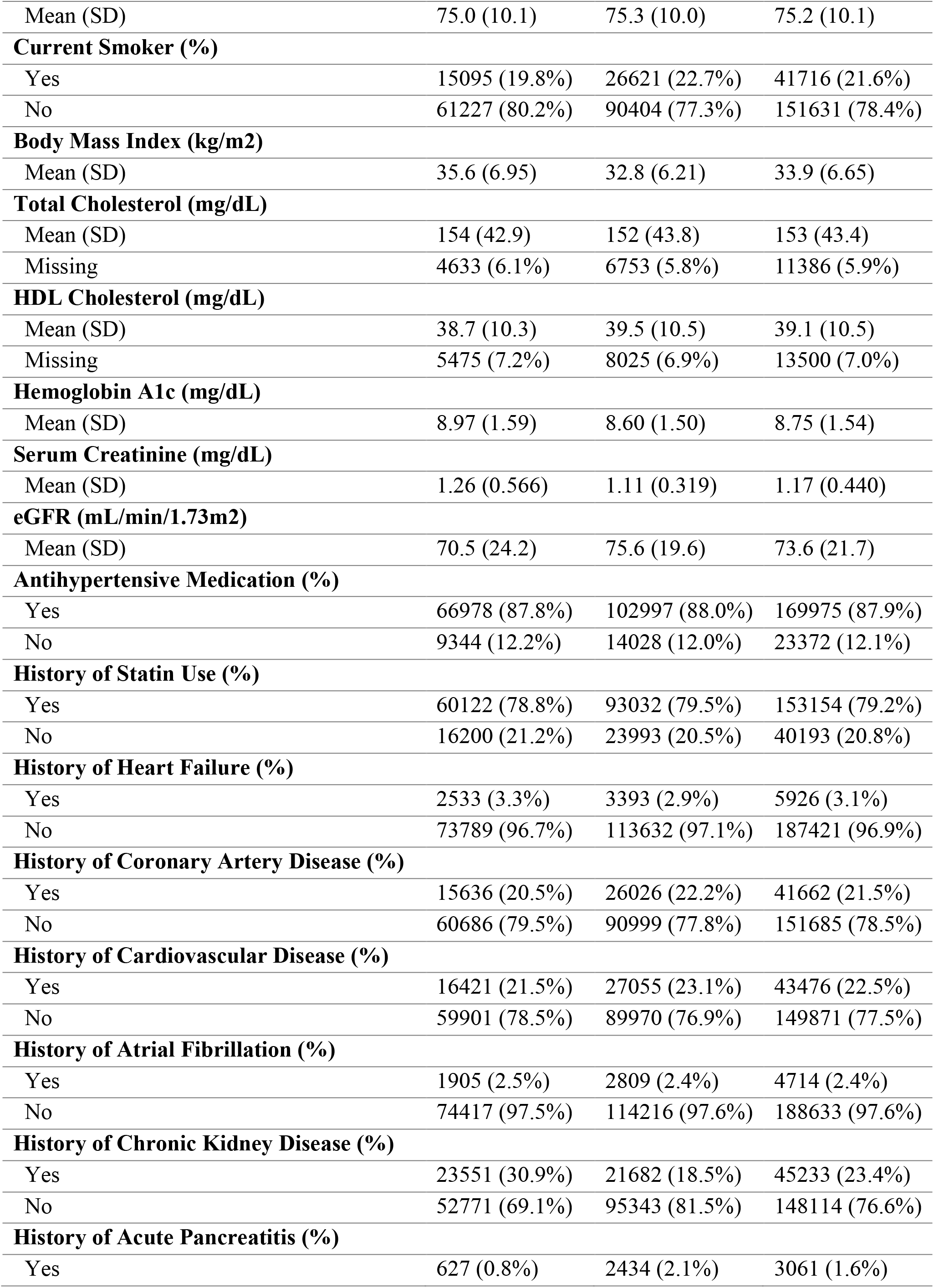
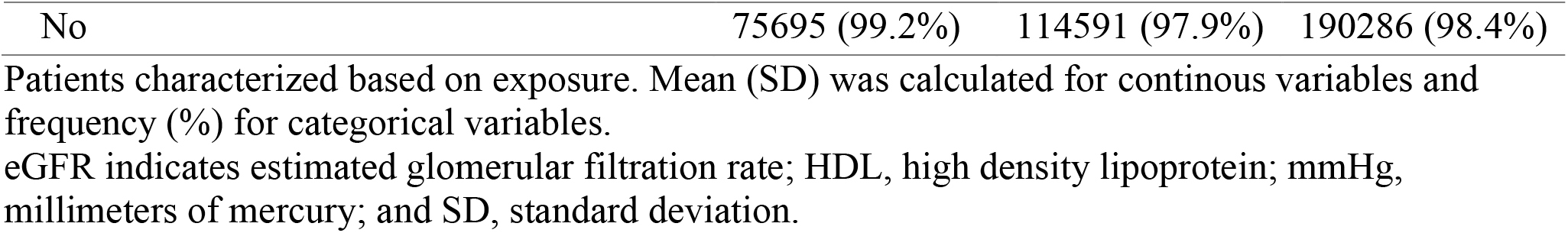
Baseline characteristics for cohort GLP1RA and SGLT2i users.

|  | **GLP1RA**<br>(N=76322) | **SGLT2i**<br>(N=117025) | **Overall**<br>(N=193347) |
| --- | --- | --- | --- |
| **Age (years)** |  |  |  |
| Mean (SD) | 66.4 (9.36) | 67.7 (8.94) | 67.2 (9.13) |
| **Gender** |  |  |  |
| Male | 71561 (93.8%) | 113098 (96.6%) | 184659 (95.5%) |
| Female | 4761 (6.2%) | 3927 (3.4%) | 8688 (4.5%) |
| **Race/Ethnicity** |  |  |  |
| White, non-Hispanic | 55593 (72.8%) | 82722 (70.7%) | 138315 (71.5%) |
| Black, non-Hispanic | 13073 (17.1%) | 20897 (17.9%) | 33970 (17.6%) |
| Hispanic | 4713 (6.2%) | 8589 (7.3%) | 13302 (6.9%) |
| American Indian/Alaska Native | 885 (1.2%) | 1206 (1.0%) | 2091 (1.1%) |
| Native Hawaiian/other Pacific Islander | 887 (1.2%) | 1328 (1.1%) | 2215 (1.1%) |
| Asian | 647 (0.8%) | 1333 (1.1%) | 1980 (1.0%) |
| Unknown | 524 (0.7%) | 950 (0.8%) | 1474 (0.8%) |
| **Systolic Blood Pressure (mmHg)** |  |  |  |
| Mean (SD) | 133 (17.6) | 134 (17.8) | 133 (17.8) |
| **Diastolic Blood Pressure (mmHg)** |  |  |  |
| Mean (SD) | 75.0 (10.1) | 75.3 (10.0) | 75.2 (10.1) |
| **Current Smoker (%)** |  |  |  |
| Yes | 15095 (19.8%) | 26621 (22.7%) | 41716 (21.6%) |
| No | 61227 (80.2%) | 90404 (77.3%) | 151631 (78.4%) |
| **Body Mass Index (kg/m2)** |  |  |  |
| Mean (SD) | 35.6 (6.95) | 32.8 (6.21) | 33.9 (6.65) |
| **Total Cholesterol (mg/dL)** |  |  |  |
| Mean (SD) | 154 (42.9) | 152 (43.8) | 153 (43.4) |
| Missing | 4633 (6.1%) | 6753 (5.8%) | 11386 (5.9%) |
| **HDL Cholesterol (mg/dL)** |  |  |  |
| Mean (SD) | 38.7 (10.3) | 39.5 (10.5) | 39.1 (10.5) |
| Missing | 5475 (7.2%) | 8025 (6.9%) | 13500 (7.0%) |
| **Hemoglobin A1c (mg/dL)** |  |  |  |
| Mean (SD) | 8.97 (1.59) | 8.60 (1.50) | 8.75 (1.54) |
| **Serum Creatinine (mg/dL)** |  |  |  |
| Mean (SD) | 1.26 (0.566) | 1.11 (0.319) | 1.17 (0.440) |
| **eGFR (mL/min/1.73m2)** |  |  |  |
| Mean (SD) | 70.5 (24.2) | 75.6 (19.6) | 73.6 (21.7) |
| **Antihypertensive Medication (%)** |  |  |  |
| Yes | 66978 (87.8%) | 102997 (88.0%) | 169975 (87.9%) |
| No | 9344 (12.2%) | 14028 (12.0%) | 23372 (12.1%) |
| **History of Statin Use (%)** |  |  |  |
| Yes | 60122 (78.8%) | 93032 (79.5%) | 153154 (79.2%) |
| No | 16200 (21.2%) | 23993 (20.5%) | 40193 (20.8%) |
| **History of Heart Failure (%)** |  |  |  |
| Yes | 2533 (3.3%) | 3393 (2.9%) | 5926 (3.1%) |
| No | 73789 (96.7%) | 113632 (97.1%) | 187421 (96.9%) |
| **History of Coronary Artery Disease (%)** |  |  |  |
| Yes | 15636 (20.5%) | 26026 (22.2%) | 41662 (21.5%) |
| No | 60686 (79.5%) | 90999 (77.8%) | 151685 (78.5%) |
| **History of Cardiovascular Disease (%)** |  |  |  |
| Yes | 16421 (21.5%) | 27055 (23.1%) | 43476 (22.5%) |
| No | 59901 (78.5%) | 89970 (76.9%) | 149871 (77.5%) |
| **History of Atrial Fibrillation (%)** |  |  |  |
| Yes | 1905 (2.5%) | 2809 (2.4%) | 4714 (2.4%) |
| No | 74417 (97.5%) | 114216 (97.6%) | 188633 (97.6%) |
| **History of Chronic Kidney Disease (%)** |  |  |  |
| Yes | 23551 (30.9%) | 21682 (18.5%) | 45233 (23.4%) |
| No | 52771 (69.1%) | 95343 (81.5%) | 148114 (76.6%) |
| **History of Acute Pancreatitis (%)** |  |  |  |
| Yes | 627 (0.8%) | 2434 (2.1%) | 3061 (1.6%) |

Patients characterized based on exposure. Mean (SD) was calculated for continuous variables and frequency (%) for categorical variables.
eGFR indicates estimated glomerular filtration rate; HDL, high density lipoprotein; mmHg, millimeters of mercury; and SD, standard deviation.

GLP-1RA users had reduced stroke risk compared to SGLT2i users in both unadjusted and IPW-adjusted analysis (HR = 0.67, 95%CI 0.64-0.69;HR = 0.72, 95%CI 0.69-0.75). In further analyses, we stratified patients by age, gender, race/ethnicity, BMI, HbA1c, and CKD stage (Table 2). Both younger and older Veterans experienced a greater reduction in stroke risk on GLP-1RA treatment (HR = 0.65, 95%CI 0.60 –0.70, 0.64;HR = 0.74, 95%CI 0.70 –0.77) than SGLT2i treatment. Males exhibited reduced stroke risk under GLP-1RA usage, but females had no difference in stroke risk between treatments. Stratified by race and ethnicity, GLP-1RA use was associated with lower stroke risk among White, non-Hispanics;Black, non-Hispanics, Hispanics, and Unknown. American Indian/Alaska Native, Native Hawaiian/other Pacific Islanders, and Asians did not experience a treatment-based reduction in stroke risk (HR = 0.91, 95%CI 0.63-1.31;HR = 0.73, 95%CI 0.50 - 1.06;HR = 0.65, 95%CI 0.40-1.06).

**Table 2.** Comparative stroke risk between GLP-1RA users and SGLT2i users stratified by age, gender, race/ethnicity, BMI, HbA1c, and CKD stage.

|  | HR | 95% CI |
| --- | --- | --- |
| **Age** |  |  |
| <65 years | 0.65 | [0.60 - 0.70] |
| 65+ years | 0.74 | [0.70 - 0.77] |
| **Gender** |  |  |
| Male | 0.71 | [0.69 - 0.74] |
| Female | 0.83 | [0.68 - 1.01] |
| **Race/Ethnicity** |  |  |
| White, non-Hispanics | 0.73 | [0.70 - 0.77] |
| Black, non-Hispanics | 0.70 | [0.64 - 0.76] |
| Hispanic | 0.73 | [0.62 - 0.85] |
| American Indian/Alaska Native | 0.91 | [0.63 - 1.30] |
| Native Hawaiian/Other Pacific Islander | 0.73 | [0.50 - 1.06] |
| Asian | 0.65 | [0.40 - 1.06] |
| Unknown | 0.43 | [0.22 - 0.84] |
| **BMI** |  |  |
| Underweight | 2.35 | [1.14 – 4.84] |
| Normal | 0.90 | [0.77 – 1.04] |
| Overweight | 0.75 | [0.70 - 0.81] |
| Obese | 0.69 | [0.66 - 0.72] |
| **HbA1c** |  |  |
| Normal | 0.69 | [0.38 – 1.25] |
| Pre-diabetic | 0.57 | [0.44 – 0.75] |
| Diabetic | 0.72 | [0.69 - 0.75] |
| **CKD stage** |  |  |
| Stage 1 | 0.68 | [0.63 - 0.74] |
| Stage 2 | 0.72 | [0.68 - 0.77] |
| Stage 3 | 0.70 | [0.66 - 0.75] |
| Stage 4/5 | 0.53 | [0.40 - 0.71] |

Values are presented as hazard ratios (95% confidence intervals). HRs were estimated using unweighted Cox proportional hazard models and stratified by the indicated subgroup. SGLT2i use is the reference category; HRs show risk for GLP1-RA users compared to SGLT2i users. BMI indicates body mass index; CI, confidence interval; CKD, chronic kidney disease; HbA1c, hemoglobin A1c; and HR, hazard ratio.

Analysis stratified by BMI showed overweight and obese individuals had a greater reduction of stroke risk on GLP-1RA than SGLT2i treatments (Table 2). Individuals that were underweight showed increased stroke risk with GLP-1RA usage (HR = 2.35, 95%CI 1.14 - 4.84), and individuals with a normal BMI had no comparative stroke risk reduction between treatments (HR = 0.89, 95%CI 0.77-1.04). Pre-diabetic and diabetic individuals both had reduced stroke risk on the GLP-1RA treatment, but Veterans with normal HbA1c levels did not have a difference in stroke risk between treatments. GLP1-RA users had lower risk of stroke compared to SLGT2 users across all CKD stages.

In secondary analysis, GLP-1RA users experienced reduced AIS and hemorrhagic stroke, respectively (HR = 0.61, 95%CI 0.58-0.63;HR = 0. 66, 95%CI 0.52 - 0.85). The overall HR between any stroke, ischemic stroke, and hemorrhagic stroke was found to be 0.72 (Figure 3).

**Figure 3.**
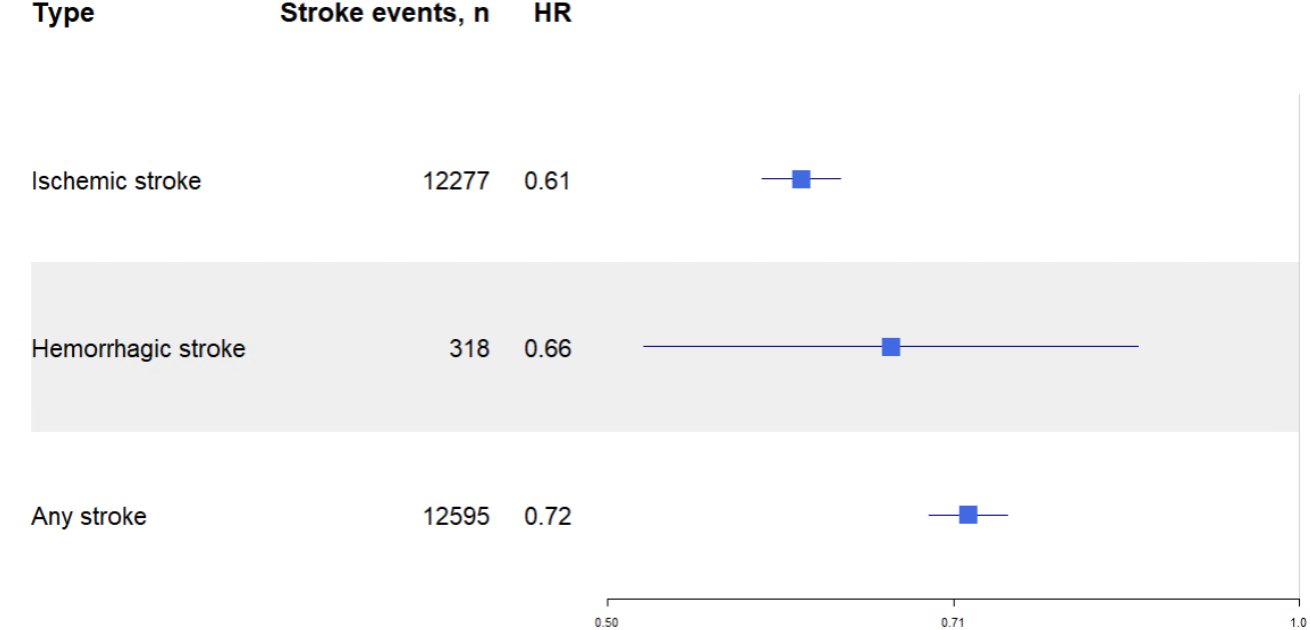
Adjusted hazard ratios for stroke sybtypes comparing between GLP-1RA and SGLT2i users. Forest plot showing hazard ratios (HRs) and 95%confidence interval (CIs) for stroke subtypes, including ischemic, hemorrhagic, and any stroke. Estimates were derived from Cox proportional hazard models using inverse probability weighting (IPW). HRs <1 indicate lower stroke risk with GLP-1RA use relative to SGLT2i use.

## Discussion and Conclusion

We found that GLP1-RA users had a lower risk of stroke than SGLT2i users. In stratified analyses, GLP-1RA users had reduced stroke risk in most subgroups, showing consistent protective effects of GLP-1RA treatment.

Analyses of comparative risks of stroke between individuals initiated on GLP-1RA and SGLT2i treatments are limited, especially of the U.S. Veteran population. Several studies have yielded similar results^6-10^. However, few papers have focused on comparing effects of the two treatments with stroke as a primary outcome. One study from Hong Kong compared incidence between GLP-1RA and SGLT2i users with stroke as their primary outcome and found reduced ischemic stroke risk only, with a comparable risk for all stroke ^6^. A smaller cohort (n=5840) based on a Hong Kong population may explain this difference in results from our secondary analyses of stroke subtype risk.

Other studies assessing differences in stroke risk between GLP-1RA and SGLT2i treatments did not separate stroke from the composite outcome. Italian, Swedish, and Scandinavian studies using a major adverse cardiovascular event composite (MACE) found similarly reduced stroke incidence of GLP-1RA users, although the GLP-1RA reduction was non-significant (HR = 1.02) in the Swedish study ^8-10^. Several studies have also found no statistically significant difference on general stroke risk between the two drug classes ^11, 12^. These differences are likely explained by differences in demographic, follow-up time, covariate adjustments, and primary outcomes between cohorts.

Following stratified analyses, females, American Indian/Alaska Native, Native Hawaiian/other Pacific Islander, Asian groups exhibited no difference in stroke risk between the two treatments. These populations were less represented in our cohort, and further research would be needed to determine if results are due to insufficient data or other confounding factors. Pre-diabetic and diabetic groups both exhibited greater stroke risk on GLP-1RA treatments, whereas those with normal HbA1c levels exhibited no preference in treatment for stroke risk reduction. This pattern is plausible: the relative stroke benefit of GLP-1RA use may be more apparent in individuals with diabetes.

When stratifying by BMI, underweight individuals were the only subgroup to have lower stroke risk reduction on GLP-1RA than SGLT2i treatment. This suggests the primary effect of GLP-1RA in stroke risk reduction may involve a reduction in BMI. Differences in biological mechanisms may help explain differences in stroke incidence between treatments. GLP-1RA focuses on stimulating insulin secretion. Common effects include lowering systolic and diastolic blood pressure, total cholesterol, improve myocardial contractility, coronary blood flow, and cardiac output ^13, 14^. This systematic reduction in inflammation and improvement in blood flow may have positive downstream effects for users. SGLT2i primarily reduce the risk of eGFR decline in patients with CKD. SGLT2i medications exhibit less effect on metabolic sites, generally resulting in less weight loss than GLP-1RA medications. With obesity being a major risk factor of stroke, GLP-1RA medications weight loss effects use may play a key role in the difference between the two medications’protective effects against stroke.

This study has some limitations. The study population is exclusive to U.S. Veterans, who may have distinct lifestyle factors. Furthermore, our cohort is predominantly male and White, non-Hispanic. Thus, generalization of our findings to other populations may be limited. As an observational cohort study, this study may also be affected by bias and causation cannot be inferred. However, propensity score matching and adjusting for covariates minimizes potential biases.

Our study has several strengths. First, the nationally integrated healthcare system data from the US Department of Veterans Affairs allows us to capture a comprehensive overview of patient clinical data. Furthermore, the large cohort size allows for adequate power to examine subgroups with sufficient data for analysis. Our further sensitivity analyses, including use of the time-varying model, allowed us to additionally confirm the robustness of our original intention-to-treat study model.

This study provides evidence of a GLP-1RA use being associated with a lower risk of stroke than SGLT2i stroke. These findings may inform future clinical decision-making and research focused on optimizing treatments strategies for patients with T2D.

## Data Availability

All relevant summary level data are included in the manuscript. Data cannot be shared publicly because of VA policies regarding data privacy and security. Data contain potentially identifying and sensitive patient information. For investigators with appropriate authorizations within the Department of Veterans Affairs, requests for data access can be made.

## Acknowledgments

We thank the Veterans Health Administration, VA Informatics and Computing Infrastructure, Centers for Medicare and Medicaid Serves, and National Death Index for providing access for the data used in this study. We appreciate the help of the data analysts who developed prior coding procedures for data extraction. We acknowledge the help of clinical staff in collecting patient data.

## Author Contributions

Sophia Sun: Conceptualization, Data Curation (lead), Formal Analysis (lead), Investigation, Methodology, Visualization, Writing –original draft

Serena Houghton: Methodology (lead), Data Curation, Formal Analysis, Investigation, Supervision, Validation

Yanping Li: Methodology, Data Curation

Xuan-Mai Nguyen: Methodology

Hugo J. Aparicio: Methodology, Supervision

Peter Wilson: Methodology, Funding Acquisition, Supervision

All authors: Writing –review and editing

## Statements and Declarations

### Ethical considerations

This study was conducted in accordance with the Declaration of Helsinki. Approval was granted by the Emory/Atlanta Veterans Affairs Medical Center (approval no. 00069355 on July 10, 2020) and the Boston Veterans Affairs Healthcare System (approval no. 1578049 on November 4, 2020) Institutional Review Boards.

### Consent to participate

The Emory/Atlanta Veterans Affairs Medical Center and the Boston Veterans Affairs Health Care System Institutional Review Boards determined that this research involved minimal risk and approved waivers for informed consent.

### Consent for publication

Not applicable.

