## Supplemental Figures for "Comparative Risk of Stroke Associated with GLP-1 Receptor Agonists and SGLT2 Inhibitors in Veterans with Type 2 Diabetes"

| Characteristic | 95% CI | HR |
|---|---|---|
| Age | | |
| <65 | (0.60 - 0.70) | 0.65 |
| 65+ | (0.70 - 0.77) | 0.74 |
| Gender | | |
| Male | (0.69 - 0.74) | 0.71 |
| Female | (0.68 - 1.01) | 0.83 |
| Race/Ethnicity | | |
| White, non-Hispanics | (0.70 - 0.77) | 0.73 |
| Black, non-Hispanics | (0.64 - 0.76) | 0.70 |
| Hispanic | (0.62 - 0.85) | 0.73 |
| American Indian/Alaska Native | (0.63 - 1.31) | 0.91 |
| Native Hawaiian/Other Pacific Islander | (0.50 - 1.06) | 0.73 |
| Asian | (0.40 - 1.06) | 0.65 |
| Unknown | (0.22 - 0.84) | 0.43 |
| BMI | | |
| Underweight | (1.14 - 4.84) | 2.35 |
| Normal | (0.77 - 1.04) | 0.89 |
| Overweight | (0.70 - 0.81) | 0.75 |
| Obese | (0.66 - 0.72) | 0.69 |
| HbA1c | | |
| Normal | (0.38 - 1.25) | 0.69 |
| Pre-diabetic | (0.44 - 0.75) | 0.57 |
| Diabetic | (0.69 - 0.75) | 0.72 |
| CKD stage | | |
| Stage 1 | (0.63 - 0.74) | 0.68 |
| Stage 2 | (0.68 - 0.77) | 0.72 |
| Stage 3 | (0.66 - 0.75) | 0.70 |
| Stage 4/5 | (0.40 - 0.71) | 0.53 |

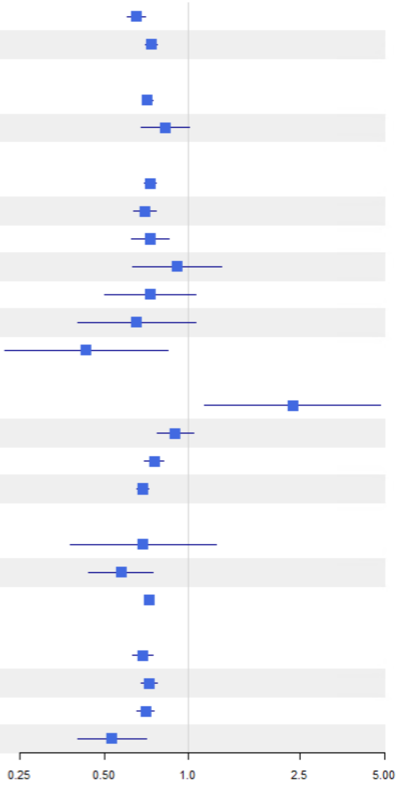


**Figure 4. Adjusted hazard ratios and confidence intervals comparing between GLP-1RA and SGLT2i users stratified by age, gender, race/ethnicity, BMI, HbA1c, and CKD stage.** Forest plot showing hazard ratios (HRs) and 95% confidence interval (CIs) stratified by different characteristics. Estimates were derived from Cox proportional hazard models using inverse probability weighting (IPW). HRs <1 indicate lower stroke risk with GLP-1RA use relative to SGLT2i use.

**Table 3. Comparative stroke risk of users on GLP-1RA or no treatment to users on SGLT2i.**

| | HR | 95% CI |
|---|---|---|
| **SGLT2i** | 1.00 | (ref) |
| **GLP-1RA** | 0.76 | (0.72-0.79) |
| **None** | 1.1 | (1.05-1.15) |

Values are presented as hazard ratios (95% confidence intervals). HRs were estimated using IPW adjusted Cox proportional hazard models. SGLT2i use is the reference category; none is classified as periods where users are on neither treatment.
CI indicates confidence interval and HR, hazard ratio.

**Table 4. Comparative stroke risk of users stratified by SGLT2i and GLP-1RA medications relative to empagliflozin use.**

| Treatment | HR | 95% CI |
|---|---|---|
| **SGLT2i** | | |
| Empagliflozin | 1.00 | (ref) |
| Canagliflozin | 0.18 | (0.8-0.41) |
| Dapagliflozin | 0.64 | (0.24-1.71) |
| Ertugliflozin | -- | -- |
| **GLP-1RA** | | |
| Albiglutide | 0.27 | (0.20-0.39) |
| Dulaglutide | 0.63 | (0.58-0.69) |
| Exenatide | 0.33 | (0.25-0.44) |
| Liraglutide | 0.53 | (0.49-0.57) |
| Lixisenatide | -- | -- |
| Semaglutide | 1.19 | (1.12-1.26) |

Values are presented as hazard ratios (95% confidence intervals). HRs were estimated using IPW weighted Cox proportional hazard models. Empagliflozin is the reference category. Ertugliflozin and Lixisenatide were dropped due to sample sizes of <10.

CI indicates confidence interval and HR, hazard ratio.
